# Association of previous medications with the risk of COVID-19: a nationwide claims-based study from South Korea

**DOI:** 10.1101/2020.05.04.20089904

**Authors:** Kyungmin Huh, Wonjun Ji, Minsun Kang, Jinwook Hong, Gi Hwan Bae, Rugyeom Lee, Yewon Na, Hyoseon Choi, Seon Yeong Gong, Jaehun Jung

## Abstract

**Background:** Identifying the association between medications taken prior to the infection of coronavirus disease (COVID-19) might be useful during the current pandemic until a proven treatment is developed. We aimed to determine whether the risk of developing COVID-19 was associated with the use of various drugs that may increase or decrease susceptibility to severe acute respiratory syndrome coronavirus 2 infection and COVID-19.

**Methods and Findings:** A case-control study was performed using a nationwide claims database of South Korea, where a large testing capacity has been available throughout the pandemic. Exposure was defined as the prescription of study drugs that would have been continued until ≤7 days before the testing for COVID-19. Adults were considered eligible if they were ≥18 years old and tested for COVID-19. Among the 65,149 eligible subjects (mean age, 48.3 years; 49.4% male), 5,172 (7.9%) were diagnosed with COVID-19. Hydroxychloroquine was not significantly associated with the risk of COVID-19 (adjusted odds ratio [aOR], 1.48; 95% CI, 0.95–2.31). In the overall population, lower risks of COVID-19 were associated with the use of camostat (aOR, 0.45; 95% CI, 0.20–1.02) and amiodarone (aOR, 0.54; 95% CI, 0.33–0.89), although the differences were not significant in the subgroup analyses. Angiotensin receptor blockers were also associated with a slightly increased risk of COVID-19 (aOR, 1.13; 95% CI, 1.01–1.26), which was also not observed in the subgroup analysis. The study limitations include potential bias regarding the controls’ characteristics, inability to determine prescription compliance, and a lack of information regarding the severity of underlying conditions.

**Conclusions:** No medications were consistently associated with increased or decreased risks of COVID-19. These findings suggest that a more cautious approach is warranted for the clinical use of re-purposed drugs until the results are available from clinical trials.

## Introduction

Coronavirus disease (COVID-19) is a novel infectious disease caused by severe acute respiratory syndrome coronavirus 2 (SARS-CoV-2) that has spread worldwide since the first reported case in late 2019. The relatively large proportion of mildly symptomatic cases and early high viral shedding has undermined the effectiveness of the classic “identify and isolate” strategy.[1] Many countries have resorted to massive social distancing to slow the spread of this disease, although a surge of patients has overwhelmed healthcare systems in several countries.

Most patients with COVID-19 have a mild course of the disease, although respiratory failure and death are more common among older people or people with underlying conditions.[2-4] Novel antiviral agents or an effective vaccine will not be available in the near future, and there is significant interest in repurposing medications that are used for other indications.[5] Hydroxychloroquine (HCQ) and lopinavir/ritonavir are drugs with promising results from preclinical studies, and some clinical data have been recently reported. Other agents are reportedly effective *in vitro* against the coronaviruses that cause severe acute respiratory syndrome or Middle East respiratory syndrome, although there is no high-quality clinical evidence to support the use of those agents for treating COVID-19.

Host cell entry of SARS-CoV-2 requires angiotensin-converting enzyme 2 (ACE2).[6] The action of ACE2 is not directly affected by ACE inhibitors (ACEIs) and angiotensin receptor blockers (ARBs), which are commonly used anti-hypertensive agents, although these drugs reportedly lead to increased expression of ACE2 in various tissues.[7, 8] Thus, there is controversy regarding whether treatment using ACEIs or ARBs might increase the host’s susceptibility to SARS-CoV-2 infection and COVID-19. Furthermore, some argue that ACEI/ARB might reduce the risk of respiratory failure, as the downregulation of ACE2 is reported to correlate with severity of lung injury.[9]

Although well-designed randomized trials are needed to address these issues, it takes time to perform those trials and report the results. Therefore, controversy persists regarding the potential harm that may be associated with use of unproven drugs for treating COVID-19. Carefully curated and reliable data from a large cohort may be a useful alternative for the time being. Thus, we conducted a case-control study to evaluate the relationships between various medications and the risk of COVID-19 using a nationwide medical insurance claim database from the Republic of Korea.

## Methods

### Data source

We extracted information from the Korean Health Insurance Review & Assessment Service (HIRA) database. The HIRA is a quasi-governmental agency that reviews all claims made to the National Health Insurance Service, which is the universal single payer for healthcare in Korea. All reimbursement claims for COVID-19 tests in suspected cases are sent for review by the HIRA (Figure 1). Claims for COVID-19-related testing are made using a special “public crisis” code (MT043), and reimbursement for confirmed cases are claimed based on the International Classification of Diseases and Related Health Problems, 10th edition (ICD-10) codes for COVID-19 (B34.2, B97.2, U18, U18.1, and U07.1). We identified all tested individuals within national health insurance services coverage who were ≥18 years old using the code MT043. Among these individuals, the cases were defined as individuals with ICD-10 codes for COVID-19 and the remaining individuals were categorized as the controls (“test-negative” controls). The last date of data entry was April 8, 2020.

This study’s retrospective protocol was approved by the institutional review board of the Gil Medical Center, Gachon University College of Medicine and Science (GFIRB2020-118).

**Figure 1.**
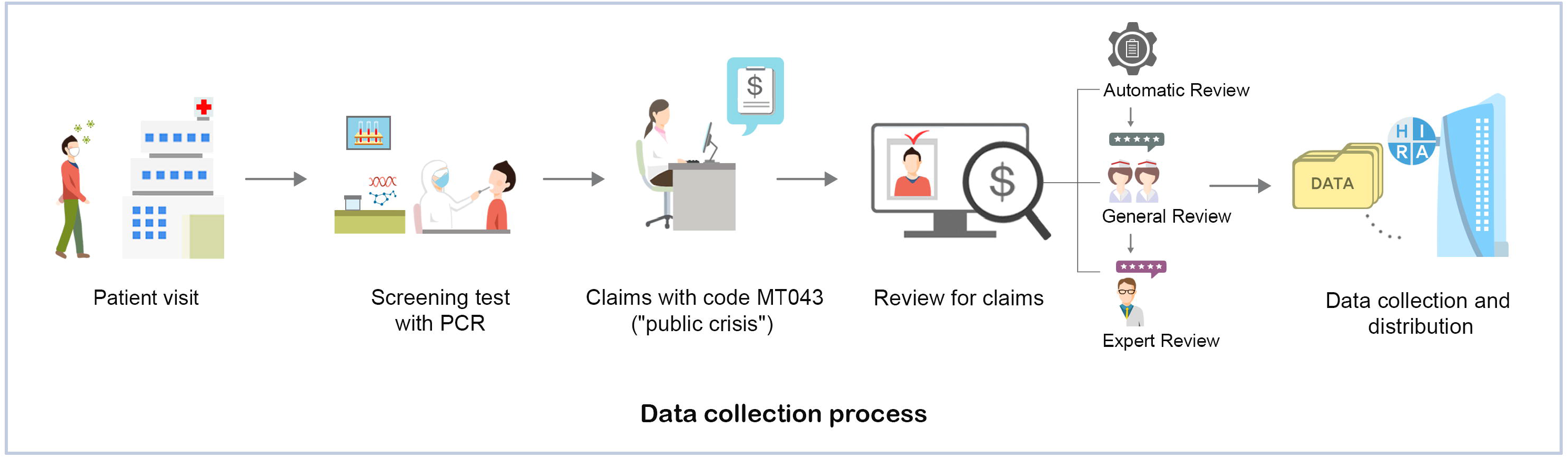
Overview of healthcare insurance claims data from Health Insurance Review & Assessment Service of Korea

### Study design and definitions

This retrospective case-control study evaluated the risk of COVID-19 according to the use of various pharmaceutical agents. Exposure was defined as the prescription of medications that would have been continued until ≤7 days before the testing for COVID-19. Drugs of interest were selected from a list of pharmaceutical agents with reported inhibitory effects against SARS-CoV-2 infection in preclinical studies, as well as agents with theoretical concerns regarding an increased risk of COVID-19 (Supplementary Table 1). Two authors (KH and WJ) reviewed the literature and selected the drugs of interest, and any disagreement was arbitrated by a third author (JJ). The maximum interval of 7 days between the prescription ending and COVID-19 testing was selected to account for the incubation period and delays in diagnosis. A preplanned subgroup analysis on the patients tested and treated in Daegu/Gyeongsangbuk-do province (DG) was conducted, as a large regional outbreak occurred in this area which led to a higher risk of community transmission.[10] Comorbidities were categorized into disease groups (Supplementary Table 2) and identified using ICD-10 codes that were entered at least twice within the 5 years before the COVID-19 tests. The Charlson comorbidity index (CCI) was calculated as previously described.[11] Healthcare utilization was evaluated based on the number of hospitalizations, the number of outpatient visits, and the number of emergency room visits within 1 year before the tests for COVID-19.

### Statistical analysis

The baseline characteristics of the cases and controls were compared using the χ^2^ test or Student’s *t* test, as appropriate. Exposures to the drugs of interest were compared using logistic regression models with sex, age, region of residence, comorbidities, healthcare utilization, and other drugs of interest as covariates. All tests were two-tailed and results were considered statistically significant at p-values of <0.05. All analyses were performed using SAS software (version 9.4; SAS Institute Inc., Cary, NC, USA).

## Results

We identified 65,149 subjects who were tested for COVID-19, including 5,172 patients (7.9%) who were diagnosed with COVID-19 (Table 1). The mean age was 48.3 years (range, 18–110 years), and 49.4% of the patients were male. The most common comorbidities were chronic lung diseases (50.9%), chronic liver diseases (44.0%), hypertension (32.8%), and diabetes (27.6%). Relative to the controls, the cases had a significantly lower mean age (44.6 years vs. 48.6 years, p<0.001) and a significantly lower proportion of male patients (44.3% vs. 48.7%, p<0.001). The controls had more comorbidities and a higher mean CCI (1.4 vs. 1.8, p<0.001), as well as more frequent hospitalizations, outpatient visits, and emergency department visits.

**Table 1.**
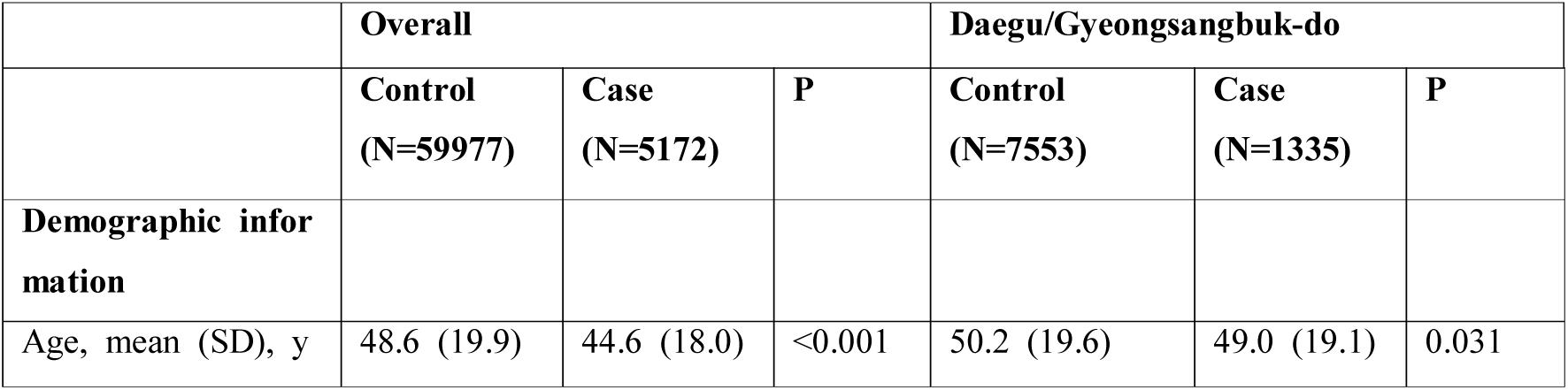
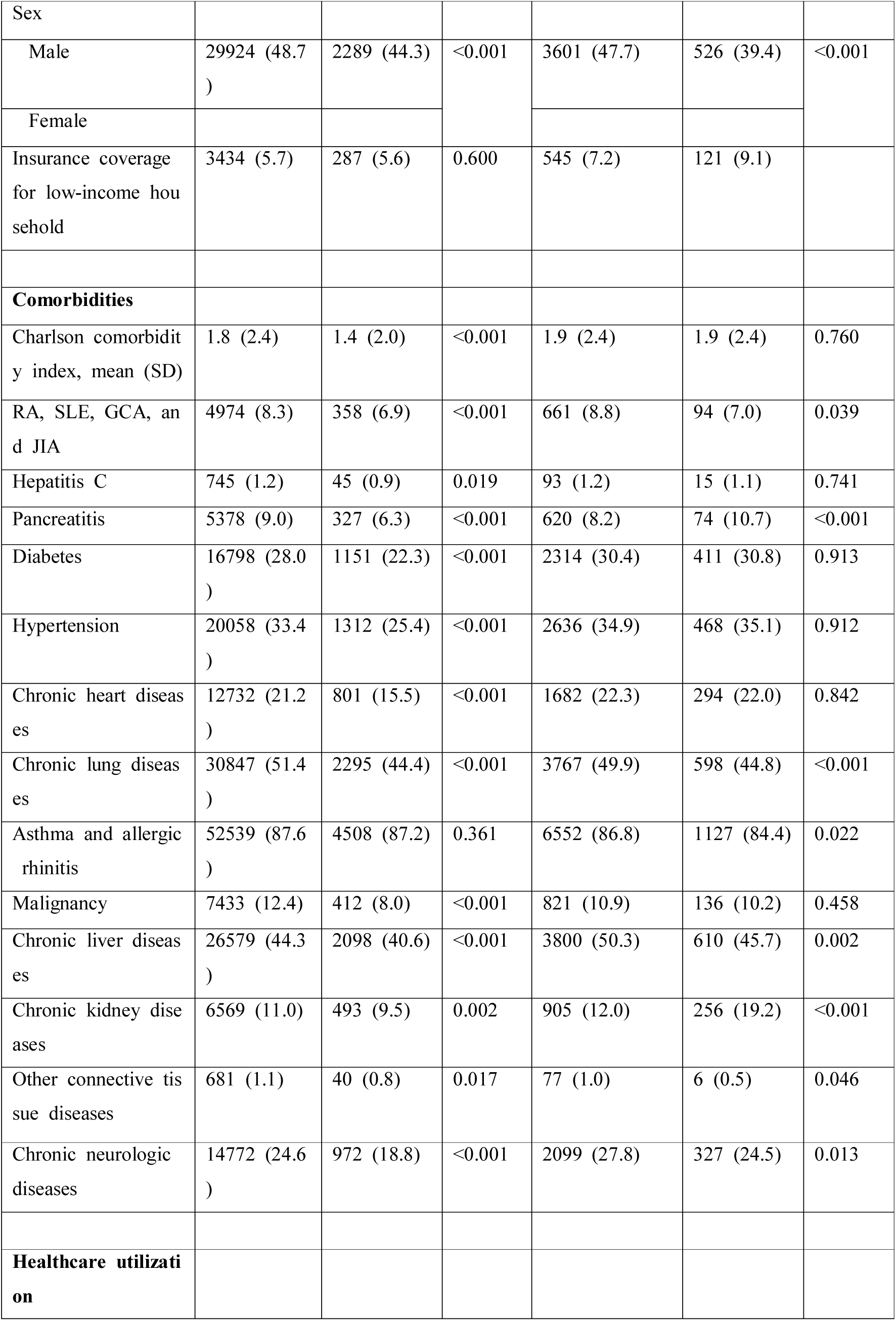
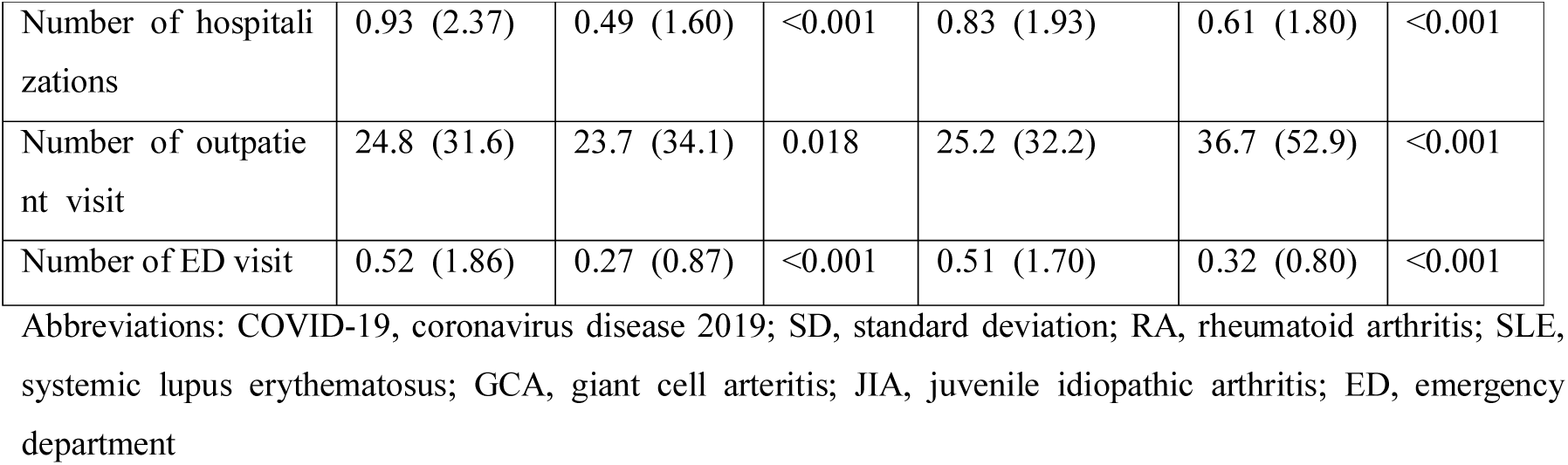
Baseline characteristics of subjects tested for COVID-19

|  | Overall |  |  | Daegu/Gyeongsangbuk-do |  |  |
| --- | --- | --- | --- | --- | --- | --- |
|  | Control<br>(N=59977) | Case<br>(N=5172) | P | Control<br>(N=7553) | Case<br>(N=1335) | P |
| **Demographic information** |  |  |  |  |  |  |
| Age, mean (SD), y | 48.6 (19.9) | 44.6 (18.0) | <0.001 | 50.2 (19.6) | 49.0 (19.1) | 0.031 |
| Sex |  |  |  |  |  |  |
| Male | 29924 (48.7) | 2289 (44.3) | <0.001 | 3601 (47.7) | 526 (39.4) | <0.001 |
| Female |  |  |  |  |  |  |
| Insurance coverage for low-income household | 3434 (5.7) | 287 (5.6) | 0.600 | 545 (7.2) | 121 (9.1) |  |
| **Comorbidities** |  |  |  |  |  |  |
| Charlson comorbidity index, mean (SD) | 1.8 (2.4) | 1.4 (2.0) | <0.001 | 1.9 (2.4) | 1.9 (2.4) | 0.760 |
| RA, SLE, GCA, and JIA | 4974 (8.3) | 358 (6.9) | <0.001 | 661 (8.8) | 94 (7.0) | 0.039 |
| Hepatitis C | 745 (1.2) | 45 (0.9) | 0.019 | 93 (1.2) | 15 (1.1) | 0.741 |
| Pancreatitis | 5378 (9.0) | 327 (6.3) | <0.001 | 620 (8.2) | 74 (10.7) | <0.001 |
| Diabetes | 16798 (28.0) | 1151 (22.3) | <0.001 | 2314 (30.4) | 411 (30.8) | 0.913 |
| Hypertension | 20058 (33.4) | 1312 (25.4) | <0.001 | 2636 (34.9) | 468 (35.1) | 0.912 |
| Chronic heart diseases | 12732 (21.2) | 801 (15.5) | <0.001 | 1682 (22.3) | 294 (22.0) | 0.842 |
| Chronic lung diseases | 30847 (51.4) | 2295 (44.4) | <0.001 | 3767 (49.9) | 598 (44.8) | <0.001 |
| Asthma and allergic rhinitis | 52539 (87.6) | 4508 (87.2) | 0.361 | 6552 (86.8) | 1127 (84.4) | 0.022 |
| Malignancy | 7433 (12.4) | 412 (8.0) | <0.001 | 821 (10.9) | 136 (10.2) | 0.458 |
| Chronic liver diseases | 26579 (44.3) | 2098 (40.6) | <0.001 | 3800 (50.3) | 610 (45.7) | 0.002 |
| Chronic kidney diseases | 6569 (11.0) | 493 (9.5) | 0.002 | 905 (12.0) | 256 (19.2) | <0.001 |
| Other connective tissue diseases | 681 (1.1) | 40 (0.8) | 0.017 | 77 (1.0) | 6 (0.5) | 0.046 |
| Chronic neurologic diseases | 14772 (24.6) | 972 (18.8) | <0.001 | 2099 (27.8) | 327 (24.5) | 0.013 |
| **Healthcare utilization** |  |  |  |  |  |  |
| Number of hospitalizations | 0.93 (2.37) | 0.49 (1.60) | <0.001 | 0.83 (1.93) | 0.61 (1.80) | <0.001 |
| Number of outpatient visit | 24.8 (31.6) | 23.7 (34.1) | 0.018 | 25.2 (32.2) | 36.7 (52.9) | <0.001 |
| Number of ED visit | 0.52 (1.86) | 0.27 (0.87) | <0.001 | 0.51 (1.70) | 0.32 (0.80) | <0.001 |

Abbreviations: COVID-19, coronavirus disease 2019; SD, standard deviation; RA, rheumatoid arthritis; SLE, systemic lupus erythematosus; GCA, giant cell arteritis; JIA, juvenile idiopathic arthritis; ED, emergency department

A total of 8,888 subjects were tested in DG province, where a regional outbreak occurred, and the test positivity rate in this subgroup was 15.0% (n=1,335). Relative to the overall population, the cases’ and controls’ baseline characteristics were more similar in the DG province subgroup. There were no significant differences between the DG cases and controls in terms of mean CCI (1.9 vs. 1.9, p=0.760) and the prevalences of diabetes, hypertension, chronic heart diseases, and malignancy. However, the controls still had higher proportions of chronic lung diseases (44.8% vs. 49.9%, p<0.001), chronic liver diseases (45.7% vs. 50.3%, p=0.002), and chronic neurological diseases (24.5% vs 27.8%, p=0.013). The cases had a higher rate of chronic kidney diseases (19.2% vs. 12.0%, p<0.001).

### Drugs with potential effect against SARS-CoV-2

The following drugs were included after a literature review: HCQ, camostat, statins, sirolimus, mycophenolate, amiodarone, and direct antiviral agents for hepatitis C (HCV DAA). We did not include lopinavir/ritonavir and other protease inhibitors, as the expected number of HIV cases would be too small to accurately analyze, given the prevalence of HIV infection in Korea. In the overall population, the rates of HCQ prescription were 0.44% among the cases and 0.42% among the controls (Table 2 and Supplementary Table 3). No significant association between HCQ use and COVID-19 was observed in the crude analysis (odds ratio [OR], 1.07; 95% confidence interval [CI], 0.69–1.63; p=0.774), although HCQ was marginally associated with a higher risk of COVID-19 in the multivariable analysis (OR, 1.48; 95% CI, 0.95–2.31; p=0.086). The association was more significant and the effect size was larger among subjects from DG province (Table 3). Confirmed cases of COVID-19 were more likely to have been prescribed HCQ in both the crude analysis (OR, 1.99; 95% CI, 1.08–3.67; p=0.029) and the multivariable analysis (OR, 2.53; 95% CI, 1.30–4.96; p=0.007).

**Table 2.** Risk for COVID-19 infection by previously prescribed medications in overall population

| Drug | Control<br>(N=59977) | Case<br>(N=5172) | Crude |  | Adjusted |  |
| --- | --- | --- | --- | --- | --- | --- |
|  |  |  | OR (95%<br>CI) | P | OR (95%<br>CI) | P |
| Drugs with potential therapeutic effect |  |  |  |  |  |  |
| Hydroxychloroquine | 251 (0.42) | 23 (0.44) | 1.07 (0.69-1.63) | 0.774 | 1.48 (0.95-2.31) | 0.086 |
| Camostat | 247 (0.41) | 6 (0.12) | 0.28 (0.13-0.63) | 0.002 | 0.45 (0.20-1.02) | 0.054 |
| Statins | 10413 (17.36) | 690 (13.34) | 0.73 (0.68-0.80) | <0.001 | 0.99 (0.89-1.10) | 0.827 |
| Sirolimus | 22 (0.04) | 1 (0.02) | 0.53 (0.07-3.91) | 0.536 | 1.16 (0.15-8.87) | 0.888 |
| Mycophenolate | 216 (0.36) | 13 (0.25) | 0.70 (0.40-1.22) | 0.208 | 1.00 (0.56-1.79) | >0.999 |
| Amiodarone | 697 (1.16) | 17 (0.33) | 0.28 (0.17-0.45) | <0.001 | 0.54 (0.33-0.89) | 0.015 |
| HCV DAA | 5 (0.01) | 0 (0.00) |  | 0.927 |  |  |
| Drugs with concern for increased risk |  |  |  |  |  |  |
| Angiotensin receptor blockers | 9377 (15.63) | 668 (12.92) | 0.80 (0.74-0.87) | <0.001 | 1.13 (1.01-1.26) | 0.034 |
| Angiotensin converting enzyme inhibitors | 607 (1.01) | 46 (0.89) | 0.88 (0.65-1.19) | 0.396 | 1.25 (0.91-1.71) | 0.177 |
| Metformin | 3604 (6.01) | 219 (4.23) | 0.69 (0.60-0.80) | <0.001 | 0.95 (0.81-1.11) | 0.497 |
| Thiazolidinedione | 526 (0.88) | 37 (0.72) | 0.81 (0.58-1.14) | 0.229 | 1.00 (0.70-1.41) | 0.977 |
| **Drugs with no expected benefit** |  |  |  |  |  |  |
| NSAIDs | 21193 (35.34) | 1579 (30.53) | 0.80 (0.76-0.86) | <0.001 | 0.90 (0.84-0.96) | 0.001 |
| COX2 inhibitors | 1609 (2.68) | 81 (1.57) | 0.58 (0.46-0.72) | <0.001 | 0.84 (0.66-1.06) | 0.146 |
| Aspirin | 535 (0.89) | 8 (0.15) | 0.17 (0.09-0.35) | <0.001 | 0.29 (0.14-0.58) | <0.001 |
| Systemic steroid | 11809 (19.69) | 693 (13.40) | 0.63 (0.58-0.69) | <0.001 | 0.86 (0.79-0.94) | 0.001 |
| Proton pump inhibitors | 14167 (23.62) | 660 (12.76) | 0.47 (0.44-0.51) | <0.001 | 0.62 (0.57-0.68) | <0.001 |
| N-acetylcysteine | 13078 (21.81) | 710 (13.73) | 0.57 (0.53-0.62) | <0.001 | 0.72 (0.66-0.79) | <0.001 |

Abbreviations: OR, odds ratio; CI, confidence interval; HCV, hepatitis C virus; DAA, direct antiviral agent; NSAID, non-steroid anti-inflammatory drug; COX, cyclooxygenase

**Table 3.** Risk for COVID-19 infection by previously prescribed medications in subjects tested in Daegu/Gyeongsangbuk-do.

| Drug | Control<br>(N=7553) | Case<br>(N=1335) | Crude |  | Adjusted |  |
| --- | --- | --- | --- | --- | --- | --- |
|  |  |  | OR (95%<br>CI) | P | OR (95%<br>CI) | P |
| Drugs with potential therapeutic effect |  |  |  |  |  |  |
| Hydroxychloroquine | 40 (0.53) | 14 (1.05) | 1.99 (1.08-3.67) | 0.029 | 2.53 (1.30-4.96) | 0.007 |
| Camostat | 37 (0.49) | 2 (0.15) | 0.31 (0.07-1.27) | 0.102 | 0.42 (0.10-1.83) | 0.250 |
| Statins | 1392 (18.43) | 219 (16.40) | 0.87 (0.74-1.02) | 0.077 | 0.88 (0.72-1.07) | 0.192 |
| Sirolimus | 1 (0.01) | 0 (0.00) |  | 0.970 |  |  |
| Mycophenolate | 10 (0.13) | 5 (0.37) | 2.84 (0.97-8.31) | 0.057 | 2.50 (0.74-8.44) | 0.140 |
| Amiodarone | 79 (1.05) | 9 (0.67) | 0.64 (0.32-1.28) | 0.211 | 0.89 (0.42-1.88) | 0.762 |
| HCV DAA | 1 (0.01) | 0 (0.00) |  | 0.970 |  |  |

| **Drugs with concern for increased risk** |  |  |  |  |  |  |
| --- | --- | --- | --- | --- | --- | --- |
| Angiotensin receptor blockers | 1228 (16.26) | 237 (17.75) | 1.11 (0.95-1.30) | 0.175 | 1.17 (0.95-1.44) | 0.148 |
| Angiotensin converting enzyme inhibitors | 136 (1.80) | 29 (2.17) | 1.21 (0.81-1.82) | 0.354 | 1.40 (0.88-2.22) | 0.161 |
| Metformin | 466 (6.17) | 75 (5.62) | 0.91 (0.70-1.16) | 0.437 | 1.09 (0.82-1.45) | 0.558 |
| Thiazolidinedione | 76 (1.01) | 19 (1.42) | 1.42 (0.86-2.36) | 0.173 | 1.25 (0.72-2.18) | 0.430 |
| **Drugs with no expected benefit** |  |  |  |  |  |  |
| NSAIDs | 2398 (31.75) | 360 (26.97) | 0.79 (0.70-0.90) | <0.001 | 0.90 (0.78-1.03) | 0.136 |
| COX2 inhibitors | 203 (2.69) | 25 (1.87) | 0.69 (0.46-1.05) | 0.085 | 0.74 (0.47-1.17) | 0.200 |
| Aspirin | 63 (0.83) | 2 (0.15) | 0.18 (0.04-0.73) | 0.017 | 0.24 (0.06-1.01) | 0.052 |
| Systemic steroid | 1286 (17.03) | 155 (11.61) | 0.64 (0.54-0.77) | <0.001 | 0.78 (0.64-0.94) | 0.011 |
| PPI | 1817 (24.06) | 191 (14.31) | 0.53 (0.45-0.62) | <0.001 | 0.58 (0.48-0.70) | <0.001 |
| N-acetylcysteine | 1541 (20.40) | 170 (12.73) | 0.57 (0.48-0.68) | <0.001 | 0.68 (0.57-0.82) | <0.001 |

Abbreviations: OR, odds ratio; CI, confidence interval; HCV, hepatitis C virus; DAA, direct antiviral agent; NSAID, non-steroid anti-inflammatory drug; COX, cyclooxygenase

In contrast, camostat was marginally associated with a lower risk of COVID-19 in the overall population (adjusted odds ratio [aOR], 0.45; 95% CI, 0.20–1.02; p=0.054). The confidence intervals were too broad to confirm the association in the multivariable analysis of the DG province subgroup, although the effect size remained stable. Controls were more likely to be on prescription for statins, but there was no significant difference in adjusted analyses in both populations. Sirolimus was not associated with the risk of COVID-19 in the overall population, and the sample of sirolimus users in the DG province was too small for comparison. Mycophenolate was also not associated with the risk of COVID-19 in the overall population, but was non-significantly related to an increased risk of COVID-19 in the DG province subgroup. Interestingly, amiodarone was associated with a significantly decreased risk of COVID-19 in the overall population (aOR, 0.54; 95% CI, 0.33–0.89; p=0.015), although this association was not observed in DG province (aOR, 0.89; 95% CI, 0.42–1.88; p=0.762). The use of HCV DAA was observed for 0.01% of the controls and none of the patients in the overall and DG populations, which was too small a sample for a meaningful comparison.

### Drugs with hypothetical concerns regarding an increased risk of COVID-19

After statins, ARBs were the second most commonly prescribed medications (15.4% of the subjects). Although the cases were less likely to receive ARBs (OR, 0.80; 95% CI, 0.74–0.87; p<0.001), ARB use was associated with a higher risk of COVID-19 after adjusting for comorbidities and other concomitant medications (aOR, 1.13; 95% CI, 1.01–1.26; p=0.034). However, the difference did not reach statistical significance in subjects in DG area (aOR, 1.17; 95% CI, 0.95–1.44; p=0.148). The use of ACEIs was much less common (1.0%, n=653), and ACEI use was not significantly associated with COVID-19. The controls were more likely to receive oral hypoglycemic agents (metformin and thiazolidinediones) and statins, although these drugs were not significantly associated with the risk of COVID-19 in the multivariable analyses.

## Discussion

This large case-control study evaluated Korean health insurance claims and revealed that no drugs were consistently associated with the risk for COVID-19. However, camostat was associated with a marginally reduced risk of COVID-19. In this context, SARS-CoV and SARS-CoV-2 engage ACE2 as the receptor for host cell entry, with the spike (S) protein of coronavirus binding to ACE2.[6, 12] In this process, S protein must be primed by host cell proteinases, which include a serine protease known as TMPRSS2.[13, 14] Camostat mesylate is a serine protease inhibitor that is approved for treating chronic pancreatitis in Korea and Japan. A recent *in vitro* study demonstrated that camostat, which is a known inhibitor of TMPRSS2, blocks the entry of SARS-CoV-2 into lung cells, and our results suggest that camostat might be a useful therapeutic agent for COVID-19.[6] Results from ongoing clinical trials will provide more evidence regarding its potential effect against SARS-CoV-2 infections. Nafamostat mesylate is a similar serine protease inhibitor that has also been used in an injectable formulation for pancreatitis and as a short-acting anticoagulant in Korea and Japan. Clinical trials have been proposed for critically ill patients with COVID-19 who cannot take oral medications.

Chloroquine (CQ) has been used for treating malaria since the 1940s and has also been used for the treatment of connective tissue diseases including rheumatoid arthritis and systemic lupus erythematosus (SLE). CQ reportedly inhibits the SARS-CoV-2 *in vitro* by inhibiting glycosylation of host receptors and endosomal acidification.[15, 16] In China, CQ and HCQ have been recommended for treating COVID-19 and a small trial indicated that HCQ treatment led to a faster negative conversion of viral shedding.[17, 18] However, more recent studies have suggested a lack of clinical benefit and even the possibility of poorer outcomes.[19-21] Furthermore, the interest in CQ/HCQ has raised concerns regarding cardiac toxicity and drug shortages for patients who require these drugs for other conditions.[22] CQ is not available in Korea and HCQ is used for same indications. The present study revealed that HCQ use was not associated with a lower risk of COVID-19 in the overall population, and HCQ use was actually associated with a higher risk of COVID-19 in DG province, where the cases and controls had similar proportions of comorbidities. Therefore, our results and those of the recent clinical studies suggest that HCQ does not have a significant clinical effect against SARS-CoV-2 infection, although the results from ongoing randomized controlled trials are needed clarify its effect.

Statins, which are also known as HMC-CoA reductase inhibitors, may play a therapeutic role by reducing endothelial dysfunction by affecting ACE2 expression.[23] The present study revealed that statin use was significantly less common among the cases than among the controls. However, there was no difference in the risk of COVID-19 after adjusting for demographic factors and comorbidities.

Sirolimus, mycophenolate, and amiodarone reportedly had inhibitory effects against SARS-CoV and/or MERS-CoV in preclinical studies.[24-26] However, the present study failed to detect significant protective effects for those drugs, albeit amiodarone showed a reduced risk for COVID-19 in overall population.

The ACEI and ARB inhibit the activity of ACE1, which is a homolog to ACE2 but is not used by SARS-CoV for host cell entry.[27] However, several studies have shown that ACEIs/ARBs upregulate the expression of ACE2, which is reportedly correlated with susceptibility to SARS-CoV infection *in vitro*.[7, 8, 28, 29] Thus, concerns were raised regarding the theoretical risk for COVID-19 in patients taking ACEI/ARB. In contrast, other reports have indicated that high serum levels of angiotensin II and downregulation of ACE2 expression may be related to lung injury by respiratory viral infections.[30, 31] Therefore, ACEI/ARB has a theoretical possibility to decrease the risk for severe infection by SARS-CoV-2. However, there is no evidence that ACEI/ARB are associated with clinical outcome in either way. Major societies recommended that patients continue taking those anti-hypertensives as prescribed.[27, 32] The present study is unable to support a clear conclusion regarding ACEI/ARB use, as the risk of COVID-19 was higher in the multivariable analyses for the overall population and the DG province subpopulation, although the difference was only significant in the overall population. A recent retrospective study of 1,128 patients with COVID-19 and hypertension revealed that inpatient use of ACEIs/ARBs was associated with a lower risk of mortality.[33] Considering the proposed theoretical mechanism of ACEI/ARB interacting with the pathogenesis of COVID-19, it is possible that ACEIs/ARBs have different effects during the initial SARS-CoV-2 infection and the subsequent inflammatory stage.[9] Further studies are needed to understand the risks of SARS-CoV-2 infection among patients with hypertension.

Our study has several strengths. First, South Korea rapidly established a large testing capacity and has successfully contained the outbreak, which suggests that it is unlikely that a large number of COVID-19 cases were missed. Also, we were able to use “test-negative” controls owing to the vast testing capacity. Thus, our findings might not be replicated in other countries with more limited testing capacities, where testing cannot be offered to all patients with acute respiratory infections. Second, the single-payer universal healthcare system in Korea ensures that we had reliable data regarding medication use and comorbidities. Given that there is near-complete coverage for common illnesses, only an extremely small proportion of cases would not have data regarding the drugs of interest. Last, we used the diagnosis of COVID-19 as an outcome. In contrast, most recent studies regarding medications and COVID-19 have compared the risk of a severe clinical course among COVID-19 patients.

The present study also has several limitations. First, our design was vulnerable to bias in the characteristics of “test-negative” controls. Controls were older and had more comorbidities in our study, and we suspect that older people with underlying conditions had higher chance of developing acute respiratory symptoms which led to vigorous testing. However, we conducted robust multivariable analyses and a subgroup analysis on DG area where the risk of community transmission was substantially higher. Second, the use of claims data precludes an analysis of prescription compliance or the accuracy of comorbidity diagnoses. Third, we could not obtain data regarding the severity of the underlying conditions, performance status, and socioeconomic characteristics. Last, some drugs were prescribed infrequently and the small sample sizes limited the statistical power of the related analyses.

In conclusion, we observed that no medications were consistently associated with the risk for COVID-19. Camostat and amiodarone might be associated with lower risk, which warrants further studies. However, the use of HCQ, ACEIs/ARBs, or statins was not associated with a significant change in the risk of COVID-19. Our findings suggest that repurposing these pharmaceutical agents may not provide significant clinical benefits for the patients with COVID-19. Clinical trials are needed to generate high-quality evidence regarding the efficacy of these agents in this setting.

## Data Availability

The analysis code is included as a supplement. You can use it freely.
Raw data is held by the Korea Health Insurance Review and Assessment Service. You can use it via # opendata4covid19.

https://hira-covid19.net/

## Author contributions

Drs Huh and Jung had full access to all of the data in the study and take responsibility for the integrity of the data and the accuracy of the data analysis.

Concept and design: Huh, Ji, Jung.

Acquisition, analysis or interpretation of data: Huh, Ji, Kang, Hong, Bae, Lee, Na, Choi, Gong, Jung.

Drafting of the manuscript: Huh, Ji, Jung.

Statistical analysis: Huh, Kang, Na, Choi, Jung.

## Acknowledgement

The authors appreciate healthcare professionals dedicated to treating COVID-19 patients in Korea, and the Ministry of Health and Welfare and the Health Insurance Review & Assessment Service of Korea for sharing invaluable national health insurance claims data in a prompt manner.

## Funding

This work was supported by grants from the Gachon University Gil Medical Center (grant nos. 2018-17 and 2019-11). The sponsor of the study was not involved in the study design, analysis, and interpretation of data; writing of the report; or the decision to submit the study results for publication.

**Abbreviations**
ACE2: angiotensin-converting enzyme 2
ACEIs: ACE inhibitors
aOR: adjusted odds ratio
ARBs: angiotensin receptor blockers
CCI: Charlson comorbidity index
CI: confidence interval
COVID-19: coronavirus disease
CQ: chloroquine
HCQ: hydroxychloroquine
HIRA: Health Insurance Review & Assessment Service
ICD-10: International Classification of Diseases and Related Health Problems, 10th edition
OR: odds ratio
SARS-CoV-2: severe acute respiratory syndrome coronavirus 2

---

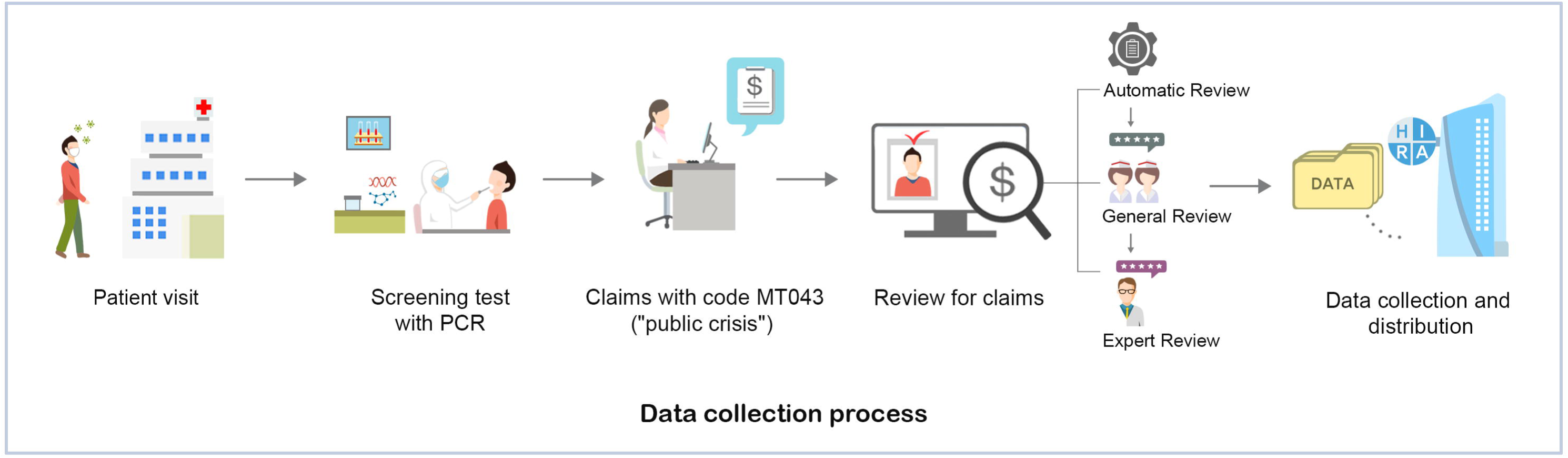

## References

1. Huh K, Shin HS, Peck KR. Emergent Strategies for the Next Phase of COVID-19. Infect Chemother. 2020;52(1):105–9. Epub 2020/02/27. doi: 10.3947/ic.2020.52.1.105. PubMed PMID: 32100487; PubMed Central PMCID: PMCPMC7113450.

2. Wu Z, McGoogan JM. Characteristics of and Important Lessons From the Coronavirus Disease 2019 (COVID-19) Outbreak in China: Summary of a Report of 72–314 Cases From the Chinese Center for Disease Control and Prevention. JAMA. 2020. doi: 10.1001/jama.2020.2648.

3. Onder G, Rezza G, Brusaferro S. Case-Fatality Rate and Characteristics of Patients Dying in Relation to COVID-19 in Italy. JAMA. 2020. doi: 10.1001/jama.2020.4683.

4. Richardson S, Hirsch JS, Narasimhan M, Crawford JM, McGinn T, Davidson KW, et al. Presenting Characteristics, Comorbidities, and Outcomes Among 5700 Patients Hospitalized With COVID-19 in the New York City Area. Jama. 2020. Epub 2020/04/23. doi: 10.1001/jama.2020.6775. PubMed PMID: 32320003.

5. Sanders JM, Monogue ML, Jodlowski TZ, Cutrell JB. Pharmacologic Treatments for Coronavirus Disease 2019 (COVID-19): A Review. JAMA. 2020. doi: 10.1001/jama.2020.6019.

6. Hoffmann M, Kleine-Weber H, Schroeder S, Krüger N, Herrler T, Erichsen S, et al. SARS-CoV-2 Cell Entry Depends on ACE2 and TMPRSS2 and Is Blocked by a Clinically Proven Protease Inhibitor. Cell. 2020;181(2):271–80.e8. Epub 2020/03/07. doi: 10.1016/j.cell.2020.02.052. PubMed PMID: 32142651; PubMed Central PMCID: PMCPMC7102627.

7. Ferrario CM, Jessup J, Chappell MC, Averill DB, Brosnihan KB, Tallant EA, et al. Effect of Angiotensin-Converting Enzyme Inhibition and Angiotensin II Receptor Blockers on Cardiac Angiotensin-Converting Enzyme 2. Circulation. 2005;111(20):2605–10. doi: doi:10.1161/CIRCULATIONAHA.104.510461.

8. Ferrario CM, Varagic J. The ANG-(1–7)/ACE2/mas axis in the regulation of nephron function. American Journal of Physiology-Renal Physiology. 2010;298(6):F1297-F305. doi: 10.1152/ajprenal.00110.2010. PubMed PMID: 20375118.

9. Sommerstein R, Kochen MM, Messerli FH, Gräni C. Coronavirus Disease 2019 (COVID-19): Do Angiotensin-Converting Enzyme Inhibitors/Angiotensin Receptor Blockers Have a Biphasic Effect? J Am Heart Assoc. 2020;9(7):e016509. Epub 2020/04/03. doi: 10.1161/jaha.120.016509. PubMed PMID: 32233753.

10. Korea Centers for Disease Control and Prevention. Updates on COVID-19 in Republic of Korea (25 March 2020). Osong, Republic of Korea: 2020.

11. Charlson ME, Pompei P, Ales KL, MacKenzie CR. A new method of classifying prognostic comorbidity in longitudinal studies: development and validation. J Chronic Dis. 1987;40(5):373–83. Epub 1987/01/01. doi: 10.1016/0021-9681(87)90171-8. PubMed PMID: 3558716.

12. Li W, Moore MJ, Vasilieva N, Sui J, Wong SK, Berne MA, et al. Angiotensin-converting enzyme 2 is a functional receptor for the SARS coronavirus. Nature. 2003;426(6965):450–4. Epub 2003/12/04. doi: 10.1038/nature02145. PubMed PMID: 14647384; PubMed Central PMCID: PMCPMC7095016.

13. Glowacka I, Bertram S, Müller MA, Allen P, Soilleux E, Pfefferle S, et al. Evidence that TMPRSS2 Activates the Severe Acute Respiratory Syndrome Coronavirus Spike Protein for Membrane Fusion and Reduces Viral Control by the Humoral Immune Response. Journal of Virology. 2011;85(9):4122–34. doi: 10.1128/jvi.02232-10.

14. Matsuyama S, Nagata N, Shirato K, Kawase M, Takeda M, Taguchi F. Efficient Activation of the Severe Acute Respiratory Syndrome Coronavirus Spike Protein by the Transmembrane Protease TMPRSS2. Journal of Virology. 2010;84(24):12658–64. doi: 10.1128/jvi.01542-10.

15. Wang M, Cao R, Zhang L, Yang X, Liu J, Xu M, et al. Remdesivir and chloroquine effectively inhibit the recently emerged novel coronavirus (2019-nCoV) in vitro. Cell Res. 2020. Epub 2020/02/06. doi: 10.1038/s41422-020-0282-0. PubMed PMID: 32020029.

16. Devaux CA, Rolain J-M, Colson P, Raoult D. New insights on the antiviral effects of chloroquine against coronavirus: what to expect for COVID-19? International Journal of Antimicrobial Agents. 2020:105938. doi: https://doi.org/10.1016/j.ijantimicag.2020.105938.

17. Gao J, Tian Z, Yang X. Breakthrough: Chloroquine phosphate has shown apparent efficacy in treatment of COVID-19 associated pneumonia in clinical studies. Biosci Trends. 2020. Epub 2020/02/20. doi: 10.5582/bst.2020.01047. PubMed PMID: 32074550.

18. Gautret P, Lagier JC, Parola P, Hoang VT, Meddeb L, Mailhe M, et al. Hydroxychloroquine and azithromycin as a treatment of COVID-19: results of an open-label non-randomized clinical trial. Int J Antimicrob Agents. 2020:105949. Epub 2020/03/25. doi: 10.1016/j.ijantimicag.2020.105949. PubMed PMID: 32205204; PubMed Central PMCID: PMCPMC7102549.

19. CHEN Jun LD, LIU Li, LIU Ping, XU Qingnian, XIA Lu, LING Yun, HUANG Dan, SONG Shuli, ZHANG Dandan, QIAN Zhiping, LI Tao, SHEN Yinzhong, LU Hongzhou. A pilot study of hydroxychloroquine in treatment of patients with common coronavirus disease-19 (COVID-19). J Zhejiang Univ (Med Sci). 2020;49(1):0-. doi: 10.3785/j.issn.1008-9292.2020.03.03.

20. Molina JM, Delaugerre C, Le Goff J, Mela-Lima B, Ponscarme D, Goldwirt L, et al. No evidence of rapid antiviral clearance or clinical benefit with the combination of hydroxychloroquine and azithromycin in patients with severe COVID-19 infection. Médecine et Maladies Infectieuses. 2020. doi: https://doi.org/10.1016/j.medmal.2020.03.006.

21. Magagnoli J, Narendran S, Pereira F, Cummings T, Hardin JW, Sutton SS, et al. Outcomes of hydroxychloroquine usage in United States veterans hospitalized with Covid-19. medRxiv. 2020:2020.04.16.20065920. doi: 10.1101/2020.04.16.20065920.

22. Yazdany J, Kim AHJ. Use of Hydroxychloroquine and Chloroquine During the COVID-19 Pandemic: What Every Clinician Should Know. Annals of Internal Medicine. 2020. doi: 10.7326/m20-1334.

23. Fedson DS, Opal SM, Rordam OM. Hiding in Plain Sight: an Approach to Treating Patients with Severe COVID-19 Infection. mBio. 2020;11(2). Epub 2020/03/22. doi: 10.1128/mBio.00398-20. PubMed PMID: 32198163; PubMed Central PMCID: PMCPMC7157814.

24. Kindrachuk J, Ork B, Hart BJ, Mazur S, Holbrook MR, Frieman MB, et al. Antiviral potential of ERK/MAPK and PI3K/AKT/mTOR signaling modulation for Middle East respiratory syndrome coronavirus infection as identified by temporal kinome analysis. Antimicrob Agents Chemother. 2015;59(2):1088–99. Epub 2014/12/10. doi: 10.1128/aac.03659-14. PubMed PMID: 25487801; PubMed Central PMCID: PMCPMC4335870.

25. Cheng KW, Cheng SC, Chen WY, Lin MH, Chuang SJ, Cheng IH, et al. Thiopurine analogs and mycophenolic acid synergistically inhibit the papain-like protease of Middle East respiratory syndrome coronavirus. Antiviral Res. 2015;115:9–16. Epub 2014/12/30. doi: 10.1016/j.antiviral.2014.12.011. PubMed PMID: 25542975; PubMed Central PMCID: PMCPMC7113672.

26. Aimo A, Baritussio A, Emdin M, Tascini C. Amiodarone as a possible therapy for coronavirus infection. Eur J Prev Cardiol. 2020:2047487320919233. Epub 2020/04/17. doi: 10.1177/2047487320919233. PubMed PMID: 32295404.

27. HFSA/ACC/AHA Statement Addresses Concerns Re: Using RAAS Antagonists in COVID-19 [25 Apr 2020]. Available from: https://www.acc.org/latest-in-cardiology/articles/2020/03/17/08/59/hfsa-acc-aha-statement-addresses-concerns-re-using-raas-antagonists-in-covid-19.

28. Hattermann K, Müller MA, Nitsche A, Wendt S, Donoso Mantke O, Niedrig M. Susceptibility of different eukaryotic cell lines to SARS-coronavirus. Arch Virol. 2005;150(5):1023–31. Epub 2005/01/13. doi: 10.1007/s00705-004-0461-1. PubMed PMID: 15645376; PubMed Central PMCID: PMCPMC7086824.

29. Mossel EC, Huang C, Narayanan K, Makino S, Tesh RB, Peters CJ. Exogenous ACE2 expression allows refractory cell lines to support severe acute respiratory syndrome coronavirus replication. J Virol. 2005;79(6):3846–50. Epub 2005/02/26. doi: 10.1128/jvi.79.6.3846-3850.2005. PubMed PMID: 15731278; PubMed Central PMCID: PMCPMC1075706.

30. Kuba K, Imai Y, Rao S, Gao H, Guo F, Guan B, et al. A crucial role of angiotensin converting enzyme 2 (ACE2) in SARS coronavirus–induced lung injury. Nature Medicine. 2005;11(8):875–9. doi: 10.1038/nm1267.

31. Gu H, Xie Z, Li T, Zhang S, Lai C, Zhu P, et al. Angiotensin-converting enzyme 2 inhibits lung injury induced by respiratory syncytial virus. Scientific Reports. 2016;6(1):19840. doi: 10.1038/srep19840.

32. Position Statement of the ESC Council on Hypertension on ACE-Inhibitors and Angiotensin Receptor Blockers [25 Apr 2020]. Available from: https://www.escardio.org/Councils/Council-on-Hypertension-(CHT)/News/position-statement-of-the-esc-council-on-hypertension-on-ace-inhibitors-and-ang.

33. Zhang P, Zhu L, Cai J, Lei F, Qin JJ, Xie J, et al. Association of Inpatient Use of Angiotensin Converting Enzyme Inhibitors and Angiotensin II Receptor Blockers with Mortality Among Patients With Hypertension Hospitalized With COVID-19. Circ Res. 2020. Epub 2020/04/18. doi: 10.1161/circresaha.120.317134. PubMed PMID: 32302265.

